# HLA-focused type 1 diabetes genetic risk prediction in populations of diverse ancestry

**DOI:** 10.1101/2025.08.07.25333167

**Authors:** Dominika A. Michalek, Courtney Tern, Catherine C. Robertson, Wei-Min Chen, Suna Onengut-Gumuscu, Stephen S. Rich

**Affiliations:** Department of Genome Sciences, University of Virginia, Charlottesville, VA, USA; Channing Division of Network Medicine, Brigham and Women’s Hospital, Boston, MA, USA; Department of Computational Medicine and Bioinformatics, University of Michigan, Ann Arbor, MI, USA

**Keywords:** genetic risk score, HLA, multi-ancestry, transferability, type 1 diabetes

## Abstract

**Aims/hypothesis:** Type 1 diabetes is characterized by the destruction of pancreatic beta cells. Genetic factors account for ~50% of the total risk, with variants in the Human Leukocyte Antigen (HLA) region contributing to half of this genetic risk, with research historically focused on populations of European ancestry. We developed HLA-focused type 1 diabetes genetic risk scores (T1D GRS_HLA_) utilizing single nucleotide polymorphisms (SNPs) or HLA alleles from four ancestry groups (Admixed African (AFR; T1D GRS_HLA-AFR_), Admixed American (AMR; T1D GRS_HLA-AMR_), European (EUR; T1D GRS_HLA-EUR_), Finnish (FIN; T1D GRS_HLA-FIN_) and across ancestry (ALL; T1D GRS_HLA-ALL_). We assessed the performance of genetic risk scores in each population to determine transferability of constructed scores.

**Methods:** A total of 41,689 samples and 13,695 SNPs in the HLA region were genotyped, with HLA alleles imputed using the HLA TAPAS multi-ethnic reference panel. Conditionally independent SNPs and HLA alleles associated with type 1 diabetes were identified in each population group to construct T1D GRS_HLA_ models. Generated T1D GRS_HLA_ models were used to predict HLA-focused type 1 diabetes genetic risk across four ancestry groups. Performance of each T1D GRS_HLA_ model was assessed using Receiver Operating Characteristic (ROC) Area Under the Curve (AUC) and compared statistically.

**Results:** Each T1D GRS_HLA_ model included a different number of conditionally independent HLA region SNPs (AFR, n = 5; AMR, n = 3; EUR, n = 38; FIN, n = 6; ALL, n = 36) and HLA alleles (AFR, n = 6; AMR, n = 5; EUR, n = 40; FIN, n = 8; ALL, n = 41). The ROC AUC values of T1D GRS_HLA_ from SNPs or HLA alleles were similar and ranged from 0.73 (T1D GRS_HLA-Allele-AMR_ applied to FIN) to 0.88 (T1D GRS_HLA-Allele-EUR_ to EUR). The ROC AUC using the combined set of conditionally independent SNPs (T1D GRS_HLA-SNP-ALL_) or HLA alleles (T1D GRS_HLA-Allele-ALL_) performed uniformly well across all ancestry groups, ranging from 0.82 to 0.88 for SNPs, and 0.80 to 0.87 for HLA alleles.

**Conclusions/interpretation:** T1D GRS_HLA_ models derived from SNPs performed equivalently to those derived from HLA alleles across ancestries. In addition, T1D GRS_HLA-SNP-ALL_ and GRS_HLA-Allele-ALL_ models had consistently high ROC AUC values when applied across ancestry groups. Larger studies in more diverse populations are needed to better assess the transferability of T1D GRS_HLA_ across ancestries.

**Research in Context:** *What is already known about this subject?:*

- The HLA region harbors 50% of the genetic risk in type 1 diabetes.
- The majority of the type 1 diabetes genetic risk scores (T1D GRS) used in the field were developed based on European ancestry data.

*What is the key question?:*

- Should genetic risk scores for type 1 diabetes be derived separately for each population, or can a score derived from multiple ancestries effectively capture the complexity of the HLA region and predict the risk of type 1 diabetes across diverse groups?

*What are the new findings?:*

- T1D GRS_HLA_ derived from SNPs performed equivalently to T1D GRS_HLA_ derived from HLA alleles across ancestries.
- T1D GRS_HLA_ derived from combined ancestry (ALL) data performed equivalent to, or better than, ancestry-specific T1D GRS_HLA_ models developed for each ancestry group.

*How might this impact on clinical practice in the foreseeable future?:*

- The implementation of a single T1D GRS_HLA-SNP-ALL_ constructed from multiple ancestral populations can be a useful tool for population screening aiming to identify individuals at risk for type 1 diabetes for intervention and prevention.

## Introduction

Genetic variation in HLA class I and HLA class II genes are integral to modulating immune responses [1, 2]. Numerous studies have identified a wide array of associations involving HLA alleles, with links to infectious diseases [3, 4], outcomes of organ transplantation [5], and risk of autoimmune diseases [6–8]. Type 1 diabetes results from the autoimmune destruction of the insulin-producing beta cells in the pancreas [9], requiring exogenous insulin for survival. Twin and family studies demonstrate that genetic factors contribute to ~50% of the risk of type 1 diabetes [10, 11], with variation in HLA class I (HLA-A, HLA-B, HLA-C) and HLA class II (HLA-DRB1, HLA-DQA1, HLA-DQB1) accounting for 30%-50% of genetic risk [12].

Epidemiologic studies suggest that the prevalence of type 1 diabetes in children is ~4/1000 in populations of European (EUR) ancestry [13], with the highest prevalence in Scandinavia (e.g., Finland [14]) as well as Kuwait [15]. Lower prevalence in children of African (AFR), Admixed American (AMR), South Asian (SAS), and East Asian (EAS) genetic ancestry has been observed; however, the SEARCH study reported an increasing prevalence of type 1 diabetes in non-EUR ancestry populations in the United States [16].

The inclusion of non-EUR populations in genetic studies has been limited [17, 18], particularly regarding type 1 diabetes [19, 20], which is relatively rare in children of other genetic ancestries. More recent studies have expanded to include diverse populations, resulting in the identification of novel loci and new risk variants in known loci [21]; however, the role of HLA genetic variation on risk of type 1 diabetes is high for EUR ancestry [22], although ancestry-specific HLA alleles and variants have been identified in non-EUR populations [21, 23–26].

A growing effort to utilize genetics in risk assessment has led to the development of genetic risk scores (GRSs). Inclusion of genetic variants weighted by their impact on risk provides a single number that can be applied for individualized risk assessment. The initial type 1 diabetes genetic risk score (T1D GRS) [27, 28] consisted of single nucleotide polymorphisms (SNPs) at both HLA and non-HLA sites in the genome. Many of the SNPs in the HLA region “tagged” known HLA alleles associated with type 1 diabetes risk. Although several forms of the T1D GRS have been proposed, the T1D GRS1 exhibited good performance in classifying individuals at increased risk of type 1 diabetes, distinguishing risk of type 1 diabetes from type 2 diabetes, and detecting those with type 2 diabetes who require insulin for glucose control [27]. The subsequent T1D GRS2 incorporated SNPs within the HLA region to estimate known heterozygote effects (e.g., the HLA-DR3/DR4 increased risk) [28]; however, T1D GRS1 and T1D GRS2 are based primarily on data from EUR ancestry. Recent work demonstrated that an African-American T1D GRS (seven SNPs) outperformed the T1D GRS1 in AFR ancestry populations and was equivalent to the T1D GRS1 in EUR ancestry populations [23]. There is a need to address the transferability of the impact of genetic variation in the HLA region on genetic risk in diverse ancestry groups.

In this study, we utilize dense genotyping in the HLA region in samples of EUR, AFR, AMR, and Finnish (FIN) collections to develop ancestry-specific T1D GRS_HLA_ and a combined T1D GRS_HLA_ based upon SNPs and their imputed HLA alleles. Each T1D GRS_HLA_ is applied to all populations to determine performance in each ancestry group and whether a combined (cosmopolitan) or series of ancestry-specific T1D GRS_HLA_ would be appropriate for global population screening. Furthermore, we aim to assess the difference between using HLA SNPs and HLA alleles to predict T1D risk, to determine whether SNPs serve as adequate proxies for HLA alleles. SNP genotyping is currently more affordable, globally more accessible, and simpler to perform than HLA typing, making it a more practical option for large-scale screening. Our findings suggest that the combined T1D GRS_HLA_ using SNPs may enable consistently high-risk prediction across populations.

## Methods and Materials

### Participants

Unrelated individuals were assembled through the Type 1 Diabetes Genetics Consortium (T1DGC) for use in genetic studies with the custom fine-mapping ImmunoChip genotyping array [29]. The dataset included 16,198 cases with type 1 diabetes and 25,491 controls, collected mainly in the US and Europe (ESM Table 1). Supergroups of genetic ancestry (EUR, AFR, AMR and EAS) were defined based on k-means clustering [21]. The FIN population was defined within the EUR supergroup using k-means clustering. Principal components (PCs) for clustering were derived from projecting samples onto the 1000 Genomes Project phase 3 Reference Panel using PLINK v1.9 [30]. The South Asian (SAS) ancestry group was separated from the AMR supergroup using multi-dimensional scaling (MDS) analysis implemented in KING v2.3.2 [31]. Together, the clustering resulted in six genetic ancestry groups (EUR, AFR, AMR, FIN, SAS, EAS). To control for population structure, within each group, PCs were generated using PLINK v1.9 [30] by performing principal component analysis (PCA) in unrelated control individuals and projecting the subjects with type 1 diabetes onto controls. In addition, samples from all genetic ancestry groups (ALL) were combined, and PCs were generated by projection of all samples onto 1000 Genomes Reference Panel. For PCA, we excluded regions of high linkage disequilibrium (LD) [32], pruned for short-range LD (r^2^ > 0.2 in 50-kb windows) and removed SNPs with minor allele frequency (MAF) ≤ 0.05. Individuals assigned to SAS and EAS were excluded from the primary analyses due to small sample size.

**Table 1.** Performance of HLA-focused T1D GRS_HLA-SNP-ALL_ and GRS_HLA-Allele-ALL_ compared to ancestry-specific T1D GRS_HLA_ in each ancestry.

Comparison of T1D $GRS_{HLA}$ constructed from SNPs in four ancestry groups.
| Ancestry | $GRS_{HLA-SNP}$ | $GRS_{HLA-SNP-ALL}$ | $AUC_{HLA-SNP}$ | $AUC_{HLA-SNP-ALL}$ | P* |
| --- | --- | --- | --- | --- | --- |
| AFR | AFR | ALL | 0.86 | 0.86 | 0.11 |
| AMR | AMR | ALL | 0.78 | 0.82 | $7.86 \times 10^{-6}$ |
| EUR | EUR | ALL | 0.87 | 0.88 | $4.73 \times 10^{-6}$ |
| FIN | FIN | ALL | 0.82 | 0.82 | 0.79 |

Comparison of T1D $GRS_{HLA}$ constructed from HLA alleles in four ancestry groups.
| Ancestry | $GRS_{HLA-Allele}$ | $GRS_{HLA-Allele-ALL}$ | $AUC_{HLA-Allele}$ | $AUC_{HLA-Allele-ALL}$ | P* |
| --- | --- | --- | --- | --- | --- |
| AFR | AFR | ALL | 0.86 | 0.86 | 0.72 |
| AMR | AMR | ALL | 0.75 | 0.80 | $2.30 \times 10^{-6}$ |
| EUR | EUR | ALL | 0.88 | 0.87 | $3.50 \times 10^{-22}$ |
| FIN | FIN | ALL | 0.82 | 0.82 | 0.19 |

$GRS_{HLA-SNP}/GRS_{HLA-Allele}$ - T1D $GRS_{HLA}$ constructed using ancestry group (AFR, AMR, EUR or FIN)
$GRS_{HLA-SNP-ALL}/GRS_{HLA-Allele-ALL}$ - T1D $GRS_{HLA}$ constructed using combined ancestry data (ALL)
$AUC_{HLA-SNP}/AUC_{HLA-Allele}$ - area under the curve for T1D $GRS_{HLA-SNP}/GRS_{HLA-Allele}$ applied in each ancestry group
$AUC_{HLA-SNP-ALL}/AUC_{HLA-Allele-ALL}$ - area under the curve for T1D $GRS_{HLA-SNP-ALL}/GRS_{HLA-Allele-ALL}$ applied in each ancestry group
\* DeLong test [35]

### T1D diagnosis – eligibility criteria

Diagnosis of T1D is critical in determining precision of genetic risk scores. In this analysis, the T1D cases were obtained from two primary sources, the Type 1 Diabetes Genetics Consortium (T1DGC) and the SEARCH for Diabetes in Youth Study. Exclusion of T1D (control samples) was based upon absence of a diagnosis and insulin use. In the T1DGC, the eligibility criteria were (a) diagnosis before 35 years of age, (b) the use of insulin within 6 months of diagnosis, and (c) continuous use of insulin without stopping for 6 months or more. If any question of diagnosis occurred, a Clinical Committee evaluated available records for decision on inclusion. In the SEARCH study, a case was defined by a subject under 20 years of age with physician-diagnosed diabetes, confirmed by medical records review and/or provider reports, an in-person visit, with follow-up laboratory testing (including C-peptide).

### Genotyping

Genotype data in the HLA region on human chromosome 6 (28Mbp - 34Mbp) were extracted from the ImmunoChip panel, with quality control performed as previously published [21]. Briefly, DNA samples were genotyped on the Illumina ImmunoChip, raw genotyping files were assembled, and genotype clusters were generated using the Illumina GeneTrain2 algorithm. Variant filters were applied to (1) re-annotate positions by aligning probe sequences to GRCh37 (hg19) and the removal of any variants with <100% match or multiple matches at different positions in the genome; (2) removal of variants with call rates <98%; (3) removal of variants with any discordance between duplicate or monozygotic twin samples, as confirmed by genotype-inferred relationships; (4) removal of variants with Mendelian inconsistencies in >1% of the informative trios or parent–offspring pairs, based on genotype-inferred relationships. X-chromosome heterozygosity and Y-chromosome missingness identified (and used to exclude) participants with apparent sex-chromosome anomalies or inconsistencies with the reported sex using KING version 2.1.3 [31]. Samples with a genotype call rate⍰<⍰98% were removed. Variants with genotype frequencies deviating from Hardy–Weinberg equilibrium (*P*⍰<⍰5⍰×⍰10^−5^) in unrelated EUR-ancestry controls were excluded before imputation.

### HLA Imputation

HLA imputation was performed using HLA-TAPAS implemented on the University of Michigan imputation server (https://imputationserver.sph.umich.edu/index.html) [33]. SNPs in the HLA region (28Mbp - 34Mbp) were used to predict classical alleles for HLA class I (HLA-*A*, -*B*, and -*C*) and HLA class II (HLA-*DQA1*, -*DQB1*, -*DRB1*, -*DPA1*, and -*DPB1*) genes with two-field accuracy. For quality control after imputation, any variant with imputation accuracy of r^2^ ≤ 0.5 and MAF ≤ 0.005 was removed from further analysis. All coordinates are reported in GRCh37.

### Association Analysis with Type 1 Diabetes to Select SNPs and HLA Alleles for T1D GRSHLA

SNPs and HLA class I and HLA class II alleles were analyzed for association with type 1 diabetes within each ancestry group (AFR, AMR, EUR, FIN) and using the combined dataset (ALL) from these groups. Logistic regression models were implemented in PLINK v.1.9, adjusting for five ancestry-specific principal components and using a minor allele count (MAC) ≥ 20 filter.

Conditional analyses were performed on SNPs and HLA alleles to identify statistically independent contributors to type 1 diabetes in each group (AFR, AMR, EUR, FIN and ALL) separately. A list of conditionally independent variants was developed by (1) including the most associated variant in the logistic regression model and (2) progressively incorporating the next most significant variants until reaching the last variant that surpassed the significance threshold. For each ancestry group, statistical significance was determined using a Bonferroni-corrected P-value threshold (alpha = 0.05) correcting for the total number of SNPs (P < 3.5 × 10^−6^) and HLA alleles (P < 3.6 × 10^−4^). For the combined dataset (ALL), statistical significance for SNPs was P < 3.4 × 10^−6^ and P < 2.8 × 10^−4^ for HLA alleles.

### HLA-focused Type 1 Diabetes Genetic Risk Score (T1D GRSHLA)

Within each ancestry group (AFR, AMR, EUR, FIN) and across-ancestry groups (ALL) HLA-focused type 1 diabetes genetic risk scores (T1D GRS_HLA_) were developed using two separate genetic markers (1) SNPs (T1D GRS_HLA-SNP_) and (2) classical HLA alleles (T1D GRS_HLA-Allele_). Within each group (AFR, AMR, EUR, FIN and ALL), to capture genetic contribution of the HLA region to type 1 diabetes, SNPs identified from conditional analysis were selected to create T1D GRS_HLA-SNP-AFR_, T1D GRS_HLA-SNP-AMR_, T1D GRS_HLA-SNP-EUR_, T1D GRS_HLA-SNP-FIN_ and T1D GRS_HLA-SNP-ALL_ and HLA alleles identified from conditional analyses were used to create T1D GRS_HLA-Allele-AFR_, T1D GRS_HLA-Allele-AMR_, T1D GRS_HLA-Allele-EUR_, T1D GRS_HLA-Allele-FIN_ and T1D GRS_HLA-Allele-ALL_. In total, five T1D GRS_HLA_ models were developed for each type of genetic marker. ESM Tables 2-6 provide list of SNPs included in each T1D GRS_HLA-SNP_ model (including information on effect allele, allele frequency and regression coefficient) and similarly, ESM Tables 7-11 provide the list of HLA alleles included in each T1D GRS_HLA-Allele_ model. The regression coefficients for each SNP (or classical HLA allele) included in the GRS_HLA_ model were used as weights for the individual SNPs (or HLA alleles). A GRS_HLA_ for each individual was calculated by summing the allele counts multiplied by their respective weight (KING v.2.3.2).

### Transferability of T1D GRSHLA in SNPs and HLA Alleles

Each T1D GRS_HLA_ model (AFR, AMR, EUR, FIN and ALL), constructed from SNPs and HLA alleles, was applied to individuals in all ancestry groups (AFR, AMR, EUR, FIN). All participants in each ancestry cohort would have five T1D GRS_HLA-SNP_ values, and five T1D GRS_HLA-Allele_ values. To determine the equivalence of T1D GRS_HLA_ risk prediction across ancestry-specific T1D GRS_HLA-SNP_ and T1D GRS_HLA-Allele_ models, Receiver Operating Characteristic (ROC) Area Under the Curve (AUC) values were computed using pROC R package [34]. The test for comparing two AUC values used the method of DeLong to determine equivalence of AUC values [35, 36].

## Results

A total of 41,689 samples and 13,695 SNPs genotyped in the HLA region were included in the study, comprising 16,198 individuals with type 1 diabetes (cases) and 25,491 individuals without type 1 diabetes (controls). For each ancestry group, 425 HLA alleles were imputed at 2-field resolution using a multi-ethnic HLA reference panel (HLA-TAPAS). SNPs were projected onto 1000 Genomes Project reference populations, and individuals were assigned to EUR (N = 33,601), AFR (N = 3,877), AMR (N = 1,084), and FIN (N = 2,804) genetic ancestry groups. Individuals assigned to SAS (N = 179) and EAS (N = 144) were excluded from the primary analyses due to small sample sizes, limiting the final sample size to 41,366 participants.

### Conditionally independent HLA region variants associated with type 1 diabetes risk

We tested SNPs and classical HLA alleles within the MHC region to identify variants that have independent effects on type 1 diabetes risk. The most significantly associated SNP with type 1 diabetes, across all ancestries and combined data, was rs9273363 (OR _AFR_ = 5.56, P_AFR_ = 1.04 × 10^−133^, OR _AMR_ = 3.72, P_AMR_ = 1.09 × 10^−38^, OR_EUR_ = 4.81, P_EUR_ = 8.36 × 10^−1464^, OR_FIN_ = 3.64, P_FIN_ = 1.19 × 10^−88^, OR_ALL_ = 4.76, P_ALL_ = 3.15 × 10^−1738^). When we tested HLA alleles for association, in AFR and AMR ancestry the most significant association with risk was with HLA-*DQA1*03:01* (OR_AFR_ = 5.45, P_AFR_ = 9.28 × 10^−116^; OR_AMR_ = 2.91, P_AMR_ = 2.44 × 10^−21^); while in EUR and FIN ancestry HLA-*DQB1*03:02* was most strongly associated with type 1 diabetes risk (OR_EUR_ = 5.33, P_EUR_ = 5.08 × 10^−1145^; OR_FIN_ = 3.91, P_FIN_ = 8.76 × 10^−76^). The most significantly associated HLA allele in the combined ancestry data was HLA-*DQB1*03:02* (OR_ALL_ = 5.13, P_ALL_ = 5.28 × 10^−1314^). In addition to identifying the most significant variant in each population, we conducted a stepwise conditional analysis to identify additional independently associated type 1 diabetes risk variants. The number of conditionally independent SNPs and HLA alleles is provided below in the GRS models and the details, including allele weights and allele frequencies are provided in ESM Tables 2-6 and ESM Tables 7-11, respectively.

### HLA-focused type 1 diabetes genetic risk scores (T1D GRSHLA)

Genetic risk scores for type 1 diabetes derived from the HLA region (T1D GRS_HLA_) were calculated using SNPs (T1D GRS_HLA-SNP_) and HLA alleles (T1D GRS_HLA-Allele_) across four ancestry populations (AFR, AMR, EUR, FIN) and combined data (ALL) using SNPs and imputed HLA alleles that were independently associated with type 1 diabetes. Of the four ancestry groups, EUR had the largest population size and the EUR-derived T1D GRS_HLA_ models contained the most SNPs (n=38) and HLA alleles (n=40), while other GRS_HLA_ models had less than 10 SNPs and HLA alleles with conditional independence of association with type 1 diabetes. The T1D GRS_HLA_ developed in AFR consists of 5 SNPs and 6 HLA alleles, in AMR of 3 SNPs and 5 HLA alleles, and in FIN of 6 SNPs and 8 HLA alleles (ESM Tables 3-6 and ESM Tables 8-11). The model constructed from combined data (ALL) was dominated by those SNPs and HLA alleles from the EUR population, and the T1D GRS_HLA_ models contained 36 SNPs and 41 HLA alleles (ESM Table 2 and ESM Table 7). Each T1D GRS_HLA_ model (AFR, AMR, EUR, FIN and ALL) was applied to all ancestry groups (AFR, AMR, EUR, FIN). We plotted the distribution of each T1D GRS_HLA_ in all ancestry groups separating T1D cases and controls. The distribution of T1D GRS_HLA_ within all populations exhibited significant overlap in those with type 1 diabetes and those without type 1 diabetes, whether using SNPs (Fig. 1a) or imputed HLA alleles (Fig. 1b). The bimodal distribution observed in the EUR and FIN populations can be explained by the presence of a substantial number of individuals carrying high-risk HLA haplotypes (HLA-DR3 and/or DR4), who exhibit the highest T1D GRS values. ESM Figure 1. shows the distribution of T1D GRS_HLA-Allele-ALL_, applied across all ancestry groups (AFR, AMR, EUR, FIN) and stratified by cases carrying high-risk HLA haplotypes (HLA-DR3 and/or DR4) and control subjects.

**Fig 1.**
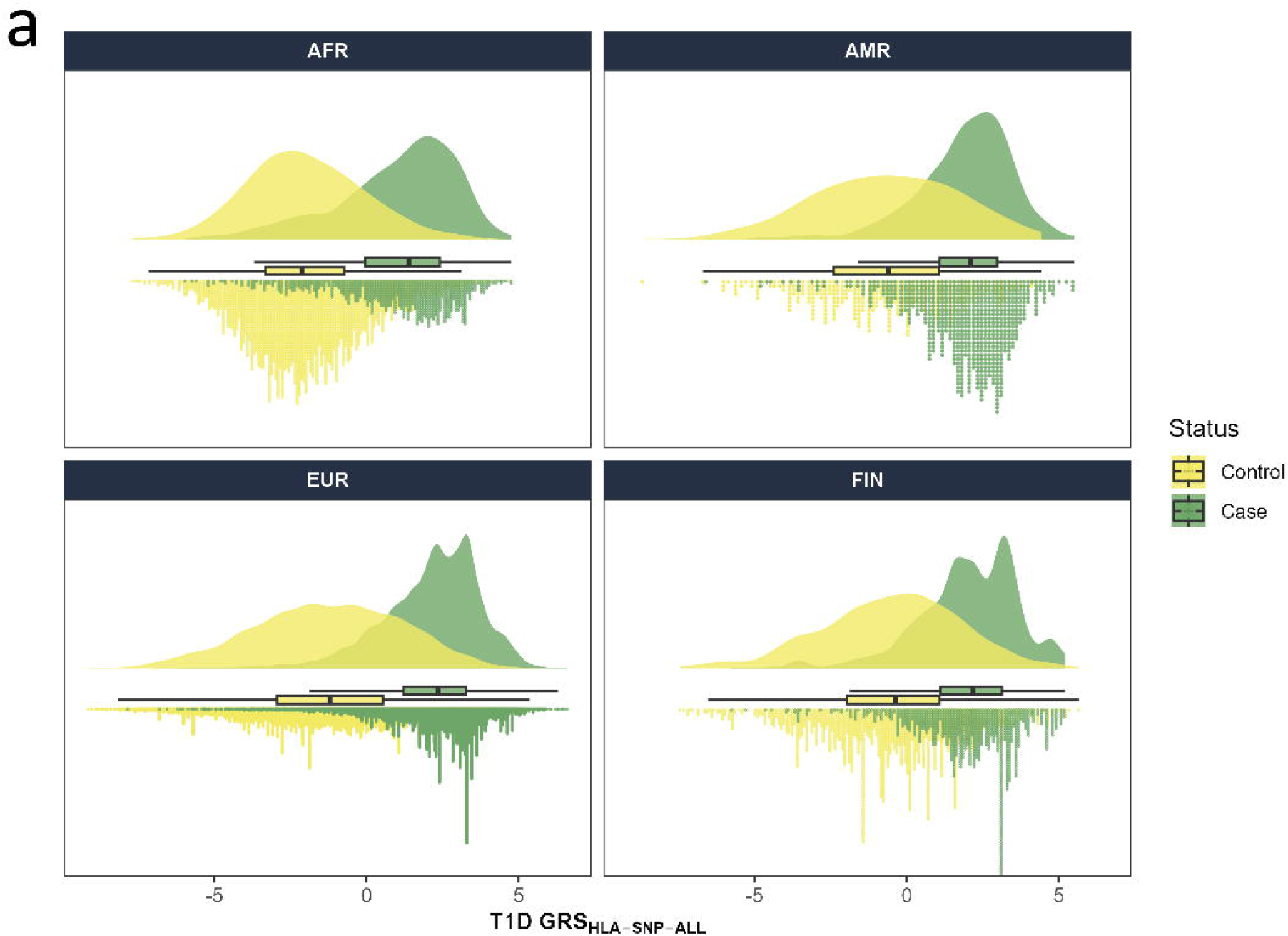
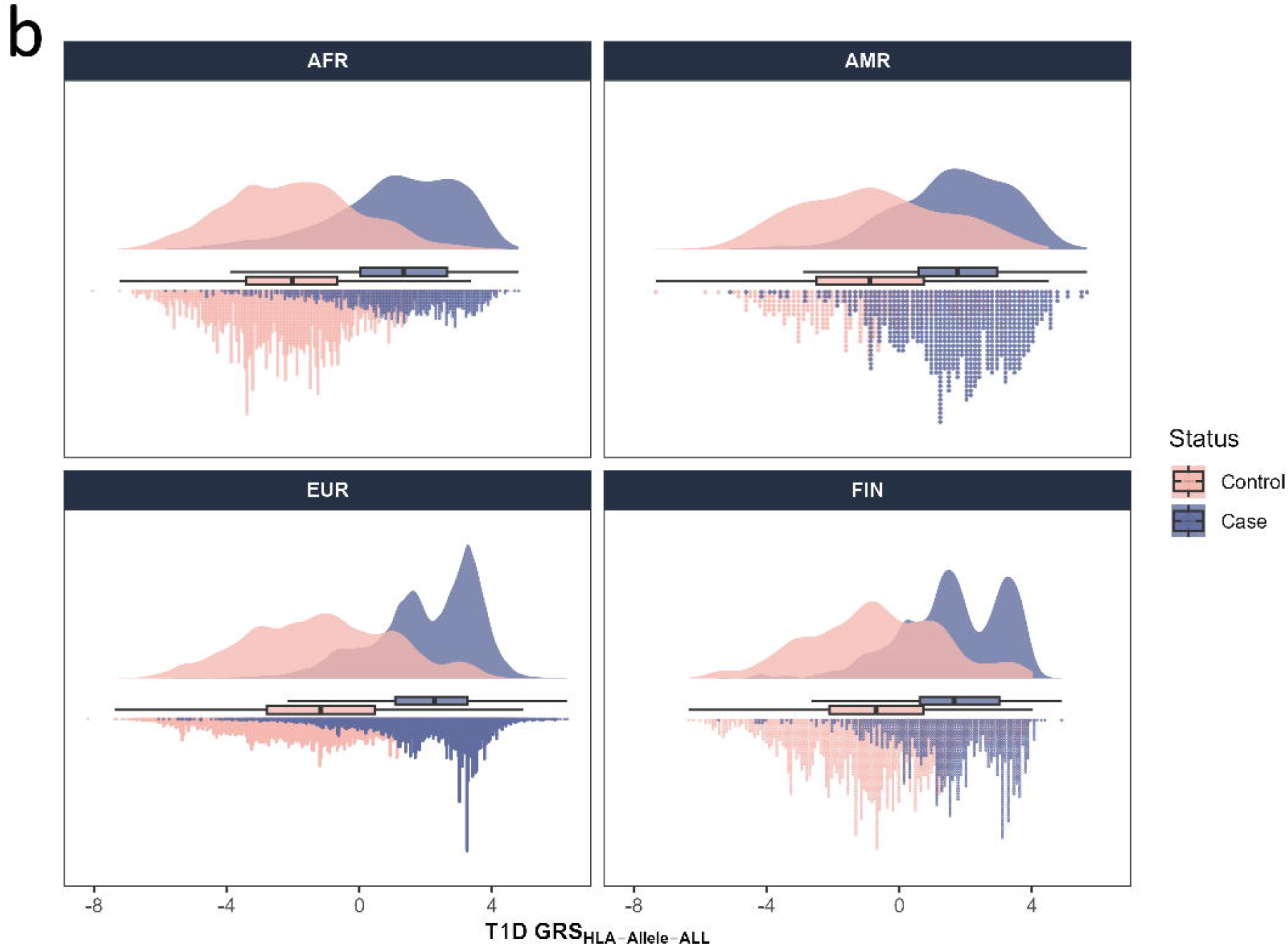
Raincloud plots of HLA-focused type 1 diabetes genetic risk scores (T1D GRS_HLA_) using combined ancestry data (ALL) SNPs (Fig. 1a) or HLA alleles (Fig. 1b) in four ancestry groups. The x-axis represents the HLA-focused T1D GRS_HLA_ by case (T1D - green (Fig. 1a), blue (Fig. 1b)) and control (no T1D - yellow (Fig. 1a), pale pink (Fig. 1b)) status in each ancestry group. The dots below the box plot represent the individual scores, while the distribution is plotted above the box plot. The box plot (median, interquartile range, and range) is shown between the upper and lower distributions.

### Prediction and transferability of type 1 diabetes using ROC curve analysis

Prediction of type 1 diabetes risk (defined by ROC AUC) using SNPs (Fig. 2a, ESM Table 12) was uniformly high, ranging from 0.74 (T1D GRS_HLA-SNP-FIN_ applied to AMR) to 0.88 (T1D GRS_HLA-SNP-ALL_ applied to EUR). Similarly, the ROC AUC using HLA alleles (Fig. 2b, ESM Table 13) ranged from 0.73 (T1D GRS_HLA-Allele-AMR_ applied to FIN) to 0.88 (T1D GRS_HLA-Allele-EUR_ to EUR). The T1D GRS_HLA_ model based upon combined data (T1D GRS_HLA-SNP-ALL_ and T1D GRS_HLA-Allele-ALL_) performed equivalently to the best individual ancestry-derived models across groups, whether using SNPs or HLA alleles. There were no significant differences in model performance, whether using SNPs or imputed HLA alleles for any comparison.

**Fig 2.**
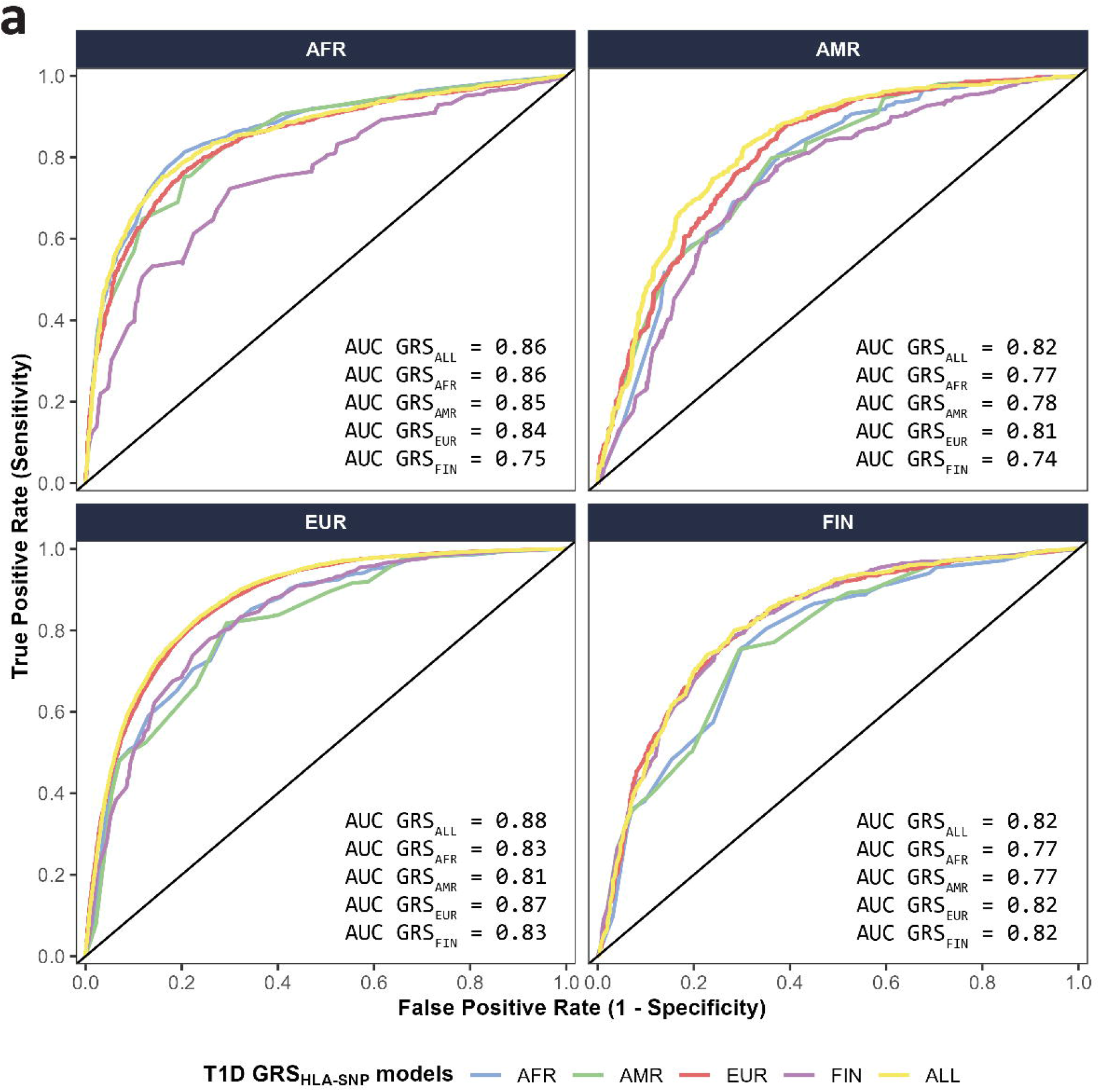
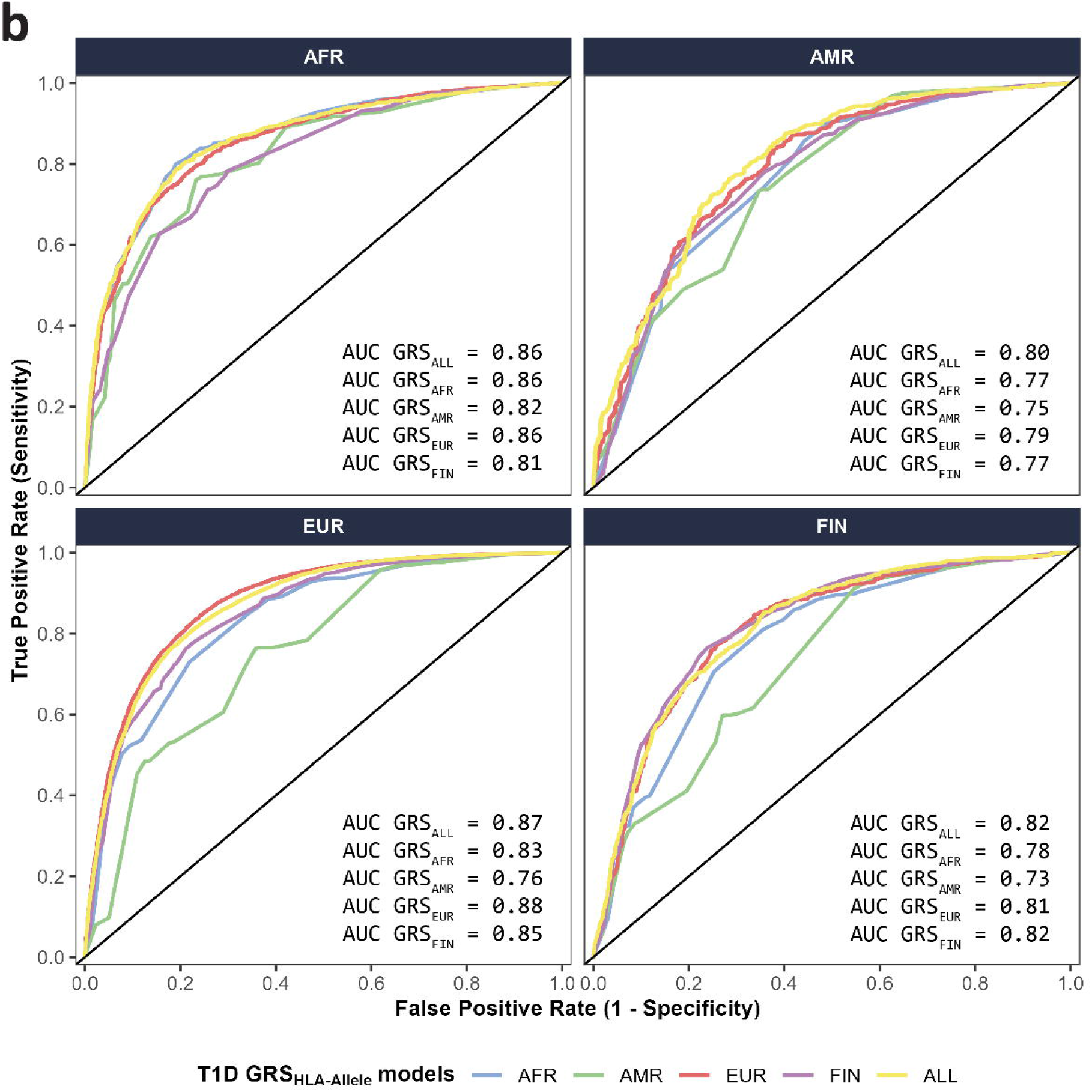
Area Under the Curve (AUC) from Receiver-Operator Characteristic (ROC) analyses of HLA-focused T1D GRS_HLA_ models based upon HLA SNPs (Fig. 2a) or HLA alleles (Fig. 2b) in four ancestry groups. Each HLA-focused T1D GRS_HLA_ model is color-coded based upon the ancestry from which it was derived (AFR - light blue, AMR - olive green, EUR - pink, FIN - light purple, ALL - yellow). Values of AUC per model are presented in the lower right corner for each ancestry group.

To evaluate transferability of scores, we compared ancestry-derived T1D GRS_HLA_ scores with T1D GRS_HLA-ALL_ scores within each ancestry group (Table 1). In AFR ancestry group, the performance of T1D GRS_HLA-SNP-ALL_ did not differ significantly from T1D GRS_HLA-SNP-AFR_ (AUC_ALL_ = 0.86 vs. AUC_AFR_ = 0.86, p = 0.11). Similarly, in FIN ancestry group, the performance of T1D GRS_HLA-SNP-ALL_ did not differ significantly from T1D GRS_HLA-SNP-FIN_ (AUC_ALL_ = 0.82 vs. AUC_FIN_ = 0.82, p = 0.79). In contrast, T1D GRS_HLA-SNP-ALL_ performed significantly better in AMR (AUC_ALL_ = 0.82 vs. AUC_AMR_ = 0.78, p = 7.86 × 10^−6^) and EUR (AUC = 0.88 vs. AUC_ALL_ = 0.87, p_EUR_ = 4.73 × 10^−6^) ancestry groups.

### T1D risk prediction in an independent cohort and the effect of non-HLA SNPs

In order to address two limitations of cross-ancestry group comparisons, we have (1) included non-HLA region SNPs [21] to T1D GRS_HLA-SNP-ALL_ model and re-calculated the T1D GRS in each ancestry and (2) conducted a validation study in a population of diverse ancestry. Incorporating non-HLA SNPs in the T1D GRS score improved prediction in all groups. In the AFR group, the AUC increased from 0.86 to 0.88, and in the AMR and FIN groups, the AUC increased from 0.82 to 0.85 and from 0.82 to 0.84, respectively. In EUR group, the AUC increased from 0.88 to 0.91. Together, these results suggest that inclusion of non-HLA SNPs increases the predictive accuracy of the T1D GRS. In a larger, genetically diverse validation cohort (510 T1D cases, 6,342 controls; 30% AFR, 18% AMR, 11% EAS, and 41% EUR), using 23 HLA-region SNPs yielded an AUC = 0.806. The inclusion of 67 non-HLA region SNPs resulted in only a slight increase in predictive performance (AUC = 0.810). Thus, the T1D GRS including all SNPs was validated with a high AUC (~0.80) even though non-HLA SNPs did not significantly improve the AUC beyond that achieved by HLA SNPs alone.

## Discussion

In this study, we constructed HLA-focused T1D Genetic Risk Scores, T1D GRS_HLA_, in different ancestry groups as well as combining data across populations. To develop genetic risk score models, we identified independently associated SNPs, HLA class I alleles, and HLA class II alleles in Admixed African (AFR), Admixed American (AMR), European (EUR), Finnish (FIN) groups and across-ancestry (ALL). We applied each T1D GRS_HLA_ model to four ancestry groups. Our results suggest that T1D GRS_HLA_ derived from combined ancestry (ALL) data performed equivalent to, or better than, ancestry-specific T1D GRS_HLA_ models defined in each ancestry group.

Each T1D GRS_HLA_ model included different numbers of conditionally independent HLA region SNPs associated with type 1 diabetes. The most significantly associated SNP with type 1 diabetes, across all ancestries and combined data, was rs9273363. This SNP has been identified as the most strongly associated with T1D risk and tags HLA-*DQB1*03:02* in European ancestry populations. As expected, HLA-*DQB1*03:02* was most strongly associated with T1D risk in the EUR and FIN groups, while in the AFR and AMR groups, the most strongly associated HLA class II allele was HLA *DQA1*03:01*, which aligns with our previous findings from recent type 1 diabetes GWAS [24]. Given the much larger EUR sample size in the GWAS (and the current HLA-focused study), the analysis of samples from all ancestry groups also identified HLA*-DQB1*03:02* as the most strongly associated HLA allele with T1D risk.

It is well known that the HLA region has undergone selective pressure [37], leading to allele frequency changes across populations. Although many of the SNPs and HLA alleles associated with T1D risk with respect to the HLA region are shared across ancestries, the size of the effect on risk can vary dramatically. In this study, the effect of HLA-*DQB1*03:02* and the tagging SNP rs9273363 on T1D risk varies by ancestry. For rs9273363, the effect size in AFR (OR = 5.56) is larger than that in AMR (OR = 3.72), EUR (OR = 4.81), and FIN (OR = 3.64). In contrast, the HLA-*DQB1*03:02* allele tagged by this SNP differs in AFR (OR = 1.68; the HLA-*DQA1*03:01* allele is most strongly associated with T1D risk, with effect OR = 5.45), AMR (not significant in this group), EUR (OR = 5.33), and FIN (OR = 3.91). These results suggest the complexity of the HLA associations with type 1 diabetes and highlight the need to develop scores that include ancestry-informed risk prediction.

Applications of genetic variation of disease in the clinic and in population screening have adopted the methodology of genetic risk scores as an approach to reduce complexity into a single metric, similar to any clinical laboratory test (e.g., HDL cholesterol) with comparison to a standard reference range. A major limitation has been the reliance on genetic data from studies of European ancestry [17]. The field has adapted T1D GRSs [27, 28] that have been generated using European-ancestry data. They perform well to distinguish type 1 diabetes from type 2 diabetes, predict progression to insulin deficiency and improve newborn screening. In addition, the T1D GRS1 has been shown to discriminate type 1 diabetes from type 2 diabetes and maturity-onset diabetes of the young (MODY) [38]. However, there is a critical need to include non-European populations in the development of more accurate genetic risk scores, and to create T1D GRS models that incorporate diverse ancestry populations. Recent applications of T1D GRS [23, 39–41] and methods development in two consortia (eMERGE and PRIMED) are beginning to address these gaps.

In our paper we are recalculating weights for associated SNPs within each ancestry group and across all individuals. This is in comparison to other methods, where weights are driven from European populations. There are increasing efforts to assess the performance of the T1D GRS1 and T1D GRS2, that were developed in EUR-ancestry populations, when applied to other populations. Recent analysis of a 67 SNP T1D GRS in an Indian population discriminated T1D cases from controls (AUC = 0.83) although lower than in EUR populations (AUC = 0.92) [42]. An updated score (T1D GRS2x) was used in the genetically diverse All of Us Research Program [43] and was highly predictive (multi-ancestry AUC = 0.860) but lower than in EUR (AUC = 0.895). Utility of multi-ancestry T1D GRSs and development across diverse ancestries show promise, yet increased sample sizes and ancestry groups will provide improvements beyond those of EUR ancestry.

This study has several strengths and limitations. Strengths of this study include use of multi-ancestry cohorts, focus on the most important genomic region for T1D risk (the HLA region), imputation to increase number of SNPs and HLA alleles (using HLA-TAPAS), and comparison of ancestry-specific SNP- and HLA allele-based genetic risk scores. However, some limitations include the smaller number of under-represented ancestry-diverse populations (AFR, AMR), and excluding potentially informative populations due to extremely small sample size (EAS, SAS). In addition, not all HLA alleles could be imputed in all populations (e.g., HLA *DQB1*02:02*). Finally, a limitation is that there is an overlap between the training data and the testing data that may affect interpretation of performance. These limitations highlight the need to expand non-EUR cohorts to better assess the prediction of ancestry-specific genetic risk scores for all diseases and, in this case, type 1 diabetes.

In summary, our data suggests that T1D GRS_HLA_ derived from SNPs performs equivalently to T1D GRS_HLA_ derived from HLA alleles across ancestries. The T1D GRS_HLA_ model derived from one ancestry is not uniformly predictive in other ancestries. Greater sample sizes in non-EUR ancestry populations are needed to develop more accurate non-EUR T1D GRS_HLA_. In addition, our results suggest that T1D GRS_HLA-ALL_ constructed from combined ancestry data performed equally well or better than ancestry-specific scores in each ancestry. While we do not have a clear explanation, we believe that the combined ancestry GRS (T1D GRS_HLA-ALL_) performed better than the individual ancestry scores, because with greater sample size we have more power to detect T1D associated SNPs. In addition, we are including individuals of diverse ancestry allowing us to tag ancestry specific T1D risk HLA alleles and haplotypes, compared to using European ancestry SNPs only.

## Abbreviations

AFR: Admixed African
ALL: All Ancestries
AMR: Admixed American
AUC: Area Under the Curve
EUR: European
FIN: Finnish
GRS: Genetic Risk Score
HLA: Human Leukocyte Antigen
MAF: Minor Allele Frequency
MODY: Maturity-Onset Diabetes of the Young
OR: Odds Ratio
PCA: Principal Component Analysis
PC: Principal Component
ROC: Receiver Operating Characteristic
SNP: Single Nucleotide Polymorphism
T1DGC: Type 1 Diabetes Genetics Consortium
T1D GRS: Type 1 Diabetes Genetic Risk Score

## Acknowledgments

The authors express their gratitude to the investigators and groups who collected and provided biological samples or data for this study, as well as to the participants whose contributions made this research possible.

## Data availability

Summary statistics are available in the NIH database for Genotype and Phenotype (dbGaP, **\*\*link will be provided\*\***), with accession number **\*\*will be provided\*\***, and the Accelerating Medicines Partnership Common Metabolic Diseases (AMP CMD) Knowledge Portal (https://hugeamp.org/). Code used to generate results is available at https://github.com/damichalek/T1DGC_GRS_HLA.

## Contribution statement

SSR and SOG conceptualized, designed the study and contributed to data acquisition. DAM, CT, CCR, WMC conducted statistical analysis. DAM, WMC, SSR and SOG were involved in data interpretation.

DAM, SSR and SOG drafted the manuscript. All authors contributed to the editing and critical revision of the manuscript. All authors read and approved the final version to be published.

The online version of this article (https://doi.org/10.1007/xxx) contains peer reviewed but unedited supplementary material.

