## Supplementary Figures for "HLA-focused type 1 diabetes genetic risk prediction in populations of diverse ancestry"

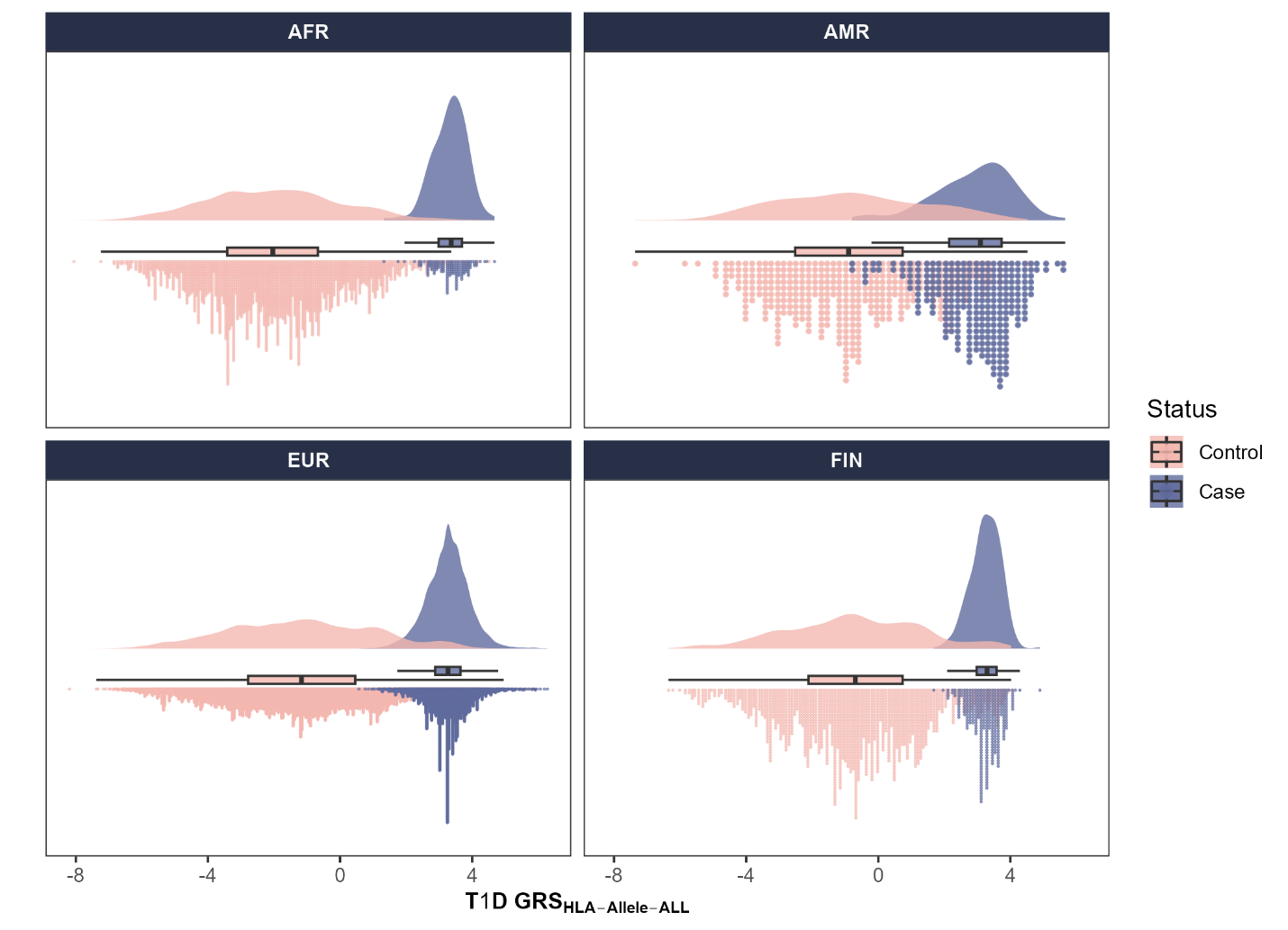

**ESM Figure 1.** Raincloud plots of HLA-focused type 1 diabetes genetic risk score, T1D GRS_HLA-Allele-ALL,_ across four ancestry groups. T1D GRS_HLA-Allele-ALL_ was derived using combined ancestry data (ALL) and then applied to each ancestry group separately. Case subjects were stratified to include only individuals carrying high-risk HLA haplotypes (e.g., HLA-DR3 and/or -DR4). The x-axis shows T1D GRS_HLA-Allele-ALL_ for type 1 diabetes cases with HLA-DR3 and/or -DR4 haplotypes (olive green) and controls without type 1 diabetes (pale pink) in each ancestry group. Each plot displays a box plot (median, interquartile range, and range). The dots below the box plot represent the individual scores, and the density distribution is plotted above the box plot.
